# Using artificial intelligence for radiotherapy clinical trial quality assurance: analysis of a multi-institutional clinical trial for neurovascular-sparing prostate stereotactic ablative radiotherapy

**DOI:** 10.64898/2026.05.27.26354252

**Authors:** Matt Doucette, Ying Zhang, Chien-Yi Liao, Mu-Han Lin, Yulong Yan, Robert T. Dess, Rahul D. Tendulkar, Aurelie Garant, Raquibul Hannan, Steve Jiang, Dan Nguyen, Neil B. Desai, Daniel X. Yang

**Author notes:** <u>Corresponding Author:</u> Daniel X. Yang, MD, Assistant Professor, Department of Radiation Oncology, UT Southwestern Medical Center, 2280 Inwood Rd, Dallas, TX 75235. NBD and DXY are co-senior authors.

## Abstract

Our study evaluated whether a deep learning auto segmentation model combined with machine learning triage can streamline radiotherapy clinical trial quality assurance (QA). We analyzed 107 stereotactic ablative radiotherapy (SABR) cases from a multi-institutional phase II clinical trial of neurovascular sparing prostate SABR, focusing on physician contours of the internal pudendal artery (IPA) as a novel organ-at-risk with substantial interobserver variability. Contours were scored by the trial principal investigator as Per-Protocol or Minor Deviation/Unacceptable. We applied a deep learning model for IPA auto-segmentation. Agreement between human and AI contours was then quantified using 14 overlap, distance, and surface metrics, and a supervised classifier was trained on these metrics to flag clinical trial protocol deviations. While AI segmentation achieved only modest geometric accuracy with mean Dice similarity coefficient of 0.446 and 95th percentile Hausdorff distance of 14.23, when incorporating all 14 metrics, a machine learning classifier yielded AUROC of 0.836, flagging all Minor Deviation/Unacceptable cases with 100% sensitivity on the 27 case hold-out set with 6 false positives and no false negatives. AI segmentation combined with metrics-based machine learning can triage protocol deviations within a multi-institution radiotherapy clinical trial, supporting prospective evaluation of AI-assisted trial QA.

## Introduction

Radiation therapy is a cornerstone of cancer treatment. The precise delivery of radiation to tumors while minimizing exposure to adjacent healthy tissue is a complex process requiring clinician expertise in target and organs-at-risk segmentation and treatment plan review. When investigating novel applications of radiotherapy, standardized guidelines for radiotherapy planning are needed within clinical trial protocols to reduce variability in radiation plan quality ^[1–3]^. While time-consuming and resource-intensive, quality assurance (QA) of radiotherapy segmentation is critical in this context. Previous analysis of radiotherapy clinical trials has demonstrated that lapses in QA and inaccurate segmentation of target volumes are associated with worse patient outcomes ^[1,4–6]^. Furthermore, adherence to protocol guidelines is important to trial conduct, as poor quality radiotherapy may also make it difficult to attribute the observed effect to the intended treatment, thereby confounding interpretability of trial results and reducing the study’s internal validity ^[2,6]^.

For patients with prostate cancer, stereotactic ablative radiotherapy (SABR), also known as stereotactic body radiotherapy (SBRT), has emerged as a standard of care curative-intent treatment option for selected patients with localized disease ^[7,8]^. While prostate SABR has been shown to achieve excellent outcomes with a favorable toxicity profile, erectile dysfunction (ED) and negative impacts on sexual quality of life domains remain major patient considerations and the most common late side effect associated with prostate radiotherapy ^[9–11]^. While the mechanism for radiotherapy-associated ED is likely multi-factorial, it may be mediated by higher dose exposure to the adjacent internal pudendal artery (IPA) and neurovascular structures. Sparing the IPA have been postulated to improve sexual function retention for prostate cancer patients undergoing SABR ^[10–12]^. This hypothesis is being prospectively tested on the recently fully accrued multi-center POTEN-C randomized clinical trial (NCT03525262) comparing novel neurovascular-sparing SABR versus standard prostate SABR for men with localized prostate cancer ^[13,14]^. This study incorporated prospective QA, including educational atlases, first case rapid-review, and centralized scoring of all submitted cases.

At the same time, artificial intelligence (AI) solutions have demonstrated high levels of auto□segmentation accuracy and efficiency, with studies demonstrating substantial reductions in contouring time while approaching inter□observer agreement levels of accuracy ^[15–20]^. Nevertheless, segmentation of novel structures such as the IPA remains challenging, as these are not typically included in standard radiotherapy planning ^[10–12]^. Our group previously developed a deep learning-model that used U-Net architecture with squeeze□and□excitation block and modality□attention to achieve promising segmentation accuracy using our institutional data ^[21]^. However, whether these algorithms remain useful within multi□institutional clinical trial settings involving heterogenous data is not yet well characterized.

Therefore, we aim to validate the previously described IPA deep-learning model using prospectively collected multi-institutional data from the POTEN-C study. Furthermore, we apply a machine learning classification model using quantitative metrics of segmentation accuracy performance to demonstrate the feasibility of using AI segmentation for automatic contour QA for patients participating in the clinical trial ^[22]^.

## Methods

### Study design and data

We conducted a retrospective, multi-institutional analysis of the POTEN-C phase II randomized trial of neurovascular-sparing SABR for localized prostate cancer ^[23]^. The POTEN-C clinical trial was approved by the Institutional Review Board at UT Southwestern under protocol STU 092017-018 and is registered on clinicaltrials.gov (NCT03525262). Written informed consent was obtained from all participants for the clinical trial. Ethical approval was provided and all methods were carried in accordance with Institutional Review Board protocol STU 082013-008 for secondary data analysis. Of 120 enrolled patients, 107 treatment plans treated 2018-2024 from seven participating centers contained planning CT, treatment planning MRI, physician-created contours of the IPA, and expert physician QA scoring for the IPA contour were included for analysis.

### AI-based IPA segmentation

Auto-segmentation of the IPA was performed with an AI model leveraging the encoder-decoder U-Net model architecture enhanced with squeeze and excite blocks and modality attention utilizing Dice similarity coefficient (DSC) and muscle saturation loss while excluding bone loss channels. Details of the model architecture have previously been described ^[21]^. CT and MRI volumes were converted from DICOM to NumPy format. CT intensities were histogram-equalized on a per-slice basis using 128 bins, and MRI volumes were min-max normalized.

When MRI was unavailable, the second input channel was zero-filled. A 128 × 128 × 64 voxel region centered on the centroid of the human-generated IPA contour was extracted from both modalities. The dual-channel input volume was passed through the trained network to produce a soft probability map of the IPA. The probability map was thresholded at 0.23 to generate a binary mask and connected components smaller than 50 voxels were removed as was performed in the previous study ^[21]^. The final mask was reinserted into the original image space and saved for evaluation.

### AI segmentation metrics

The agreement between the clinician created reference contours and the automatically generated masks was quantified with 14 metrics. The DSC, Jaccard Index (IoU), precision, sensitivity, and the Matthews correlation coefficient, were included as metrics recommended by recent critical reviews of auto□segmentation assessment in radiation oncology ^[24–29]^. Absolute and relative volume difference (AVD and RVD), mean distance□to□agreement (MDA), and both the 95th□percentile and maximum Hausdorff distances (HD□□, HD□□□),, which capture boundary discrepancies and are widely used in 3D medical imaging benchmarks, were also included ^[27,30–32]^. Moreover, surface Dice similarity coefficient at 1□ and 2□voxel tolerances (SDSC□, SDSC□) and added path length (APL_1_) at 1□voxel tolerance, which demonstrated good correlation with contour editing time, were also calculated ^[24,33]^. Metric distributions are reported as mean ± standard deviation.

### Machine learning-based identification of protocol deviation

Each patient case IPA contour previously was labeled during prospective trial QA as Per□Protocol (n = 89) or Minor Deviation/Unacceptable (n = 18) according to QA scores performed by the trial principal investigator. Five supervised learning algorithms were examined. The first was a radial□basis□function support□vector machine (SVM) ^[34]^. The second was a L2□penalized logistic regression ^[35]^. The third was an ensemble random forest model that constructs a series of decision trees each trained on a subset of the cases and metrics, and then aggregates their predictions ^[36–38]^. The fourth is an implementation of XGBoost, a commonly used tree-based machine learning method ^[39]^. The fifth is a feed□forward neural network that performs a series of weighted linear combinations of the input features followed by rectified linear unit and sigmoid activations to learn relationships across metrics that correspond to Per-Protocol vs. Minor Deviation/Unacceptable cases ^[40]^.

Class imbalance was handled with inverse□frequency weighting for the SVM and logistic regression, balanced resampling for the forest ensembles, and using the scale_pos_weight parameter for XGBoost ^[41–43]^. Model hyper□parameters were optimized via nested five□fold cross□validation ^[44]^. During five□fold cross□validation, a threshold was tuned for every model to ensure that no Minor Deviation/Unacceptable contours in the training set would be missed for human review. Final discrimination was assessed on an independent 27 case hold□out set using area under the receiver operating characteristic curve (AUROC) and confusion matrices ^[39]^.

### Statistical analysis

Models were implemented with scikit□learn 1.6.1 or the native XGBoost and Keras libraries. Group differences in geometric metrics between per protocol and deviation plans were evaluated with Welch’s *t*□tests, and two-tailed p-values were calculated ^[45,46]^. All computations were performed in Python 3.10.16 on an NVIDIA H100 GPU computing platform.

## Results

### Patient cohort

107 patients accrued to the POTEN□C clinical trial were included (Table 1). Most contours (83%, n = 89) were scored Per□Protocol, whereas 16% (n = 18) were deemed Minor Deviation or Unacceptable. Baseline clinicopathologic factors, including age at treatment (62.0 ± 15.9 vs. 61.8 ± 6.6 years) and initial PSA (6.4 ± 2.8 vs. 5.6 ± 2.3 ng/mL) were comparable between groups. Statistically significant differences were observed in human□drawn IPA volumes (16.1 ± 8.1 cm^3^ vs. 11.9 ± 8.0 cm^3^, p = 0.018) between the groups, likely reflecting the known challenge of consistently contouring vascular structures of only a few mm in diameter.^47^ However, no statistically significant difference was found between the AI-predicted IPA volumes (18.7 ± 3.0 cm^3^ vs. 17.5 ± 3.5 cm^3^, p = 0.183) and distributions of contour volume were also narrower than clinician-created contour volumes in both cohorts.

**Table 1.**
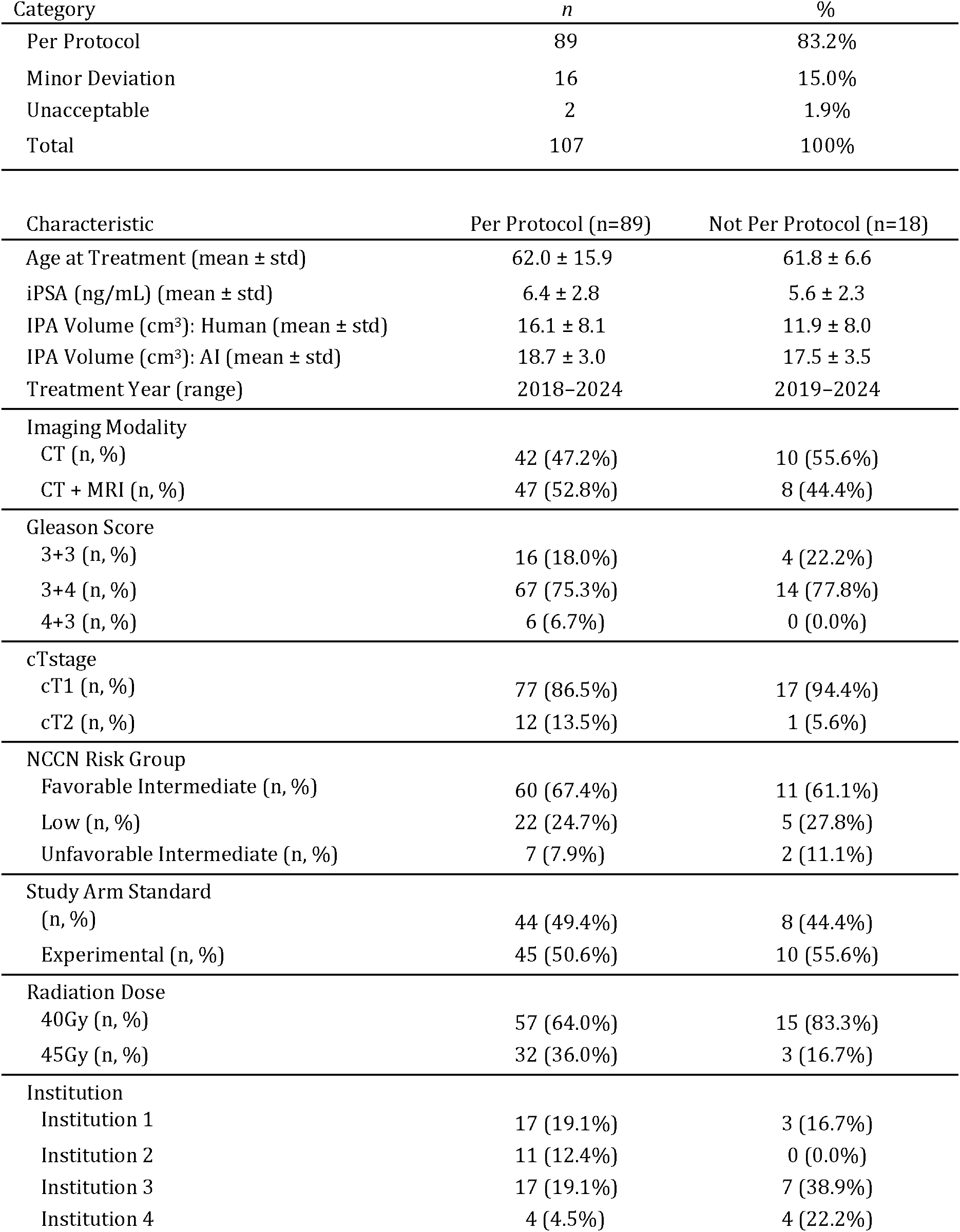
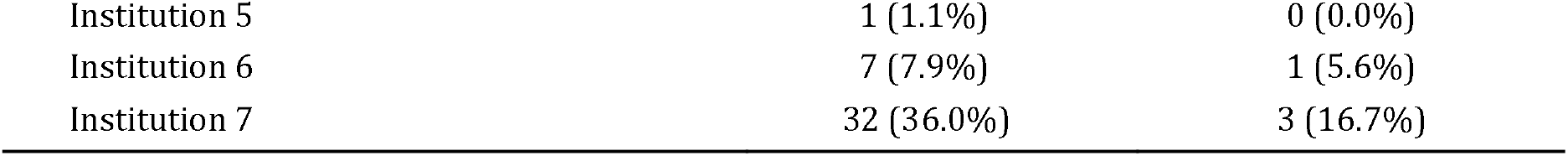
Patient and Treatment Characteristics.

### Geometric agreement between AI and reference contours

Across all cases the AI segmentation algorithm produced an average DSC of 0.446. Per□Protocol cases demonstrated significantly better correspondence between AI and human contours for multiple metrics, although the overall pattern was mixed (Table 2). The absolute volume difference (AVD) was lower by 59% (32.9 ± 32.0% vs 80.3 ± 52.9%, p = 0.001), and precision higher by 38% (0.48 ± 0.15 vs 0.35 ± 0.20, p = 0.012). The average IoU across the Per-Protocol cases was 0.313 ± 0.123, while the Minor Deviation/Unacceptable cases averaged 0.236 ± 0.149 (p = 0.050). Similarly, the Per-Protocol cases exhibited a higher mean DSC than the Minor Deviation/Unacceptable cases (0.464 ± 0.148 vs 0.358 ± 0.211, p = 0.056). No significant differences emerged for surface□based metrics (SDSC□, SDSC□) or Hausdorff distances ^[25]^.

**Table 2.** Comparison of selected metrics scores for Per Protocol and Not Per Protocol patient cases.

| Metric | Per Protocol (n=89) | Not Per Protocol (n=18) | Overall (n=107) | t | p |
| --- | --- | --- | --- | --- | --- |
| AVD | 32.96 (32.04) | 80.33 (52.9) | 40.93 (40.23) | -3.666 | *0.001 |
| Precision | 0.483 (0.149) | 0.349 (0.197) | 0.46 (0.165) | 2.728 | *0.012 |
| IoU | 0.313 (0.123) | 0.236 (0.149) | 0.3 (0.13) | 2.067 | 0.05 |
| DSC | 0.464 (0.148) | 0.358 (0.211) | 0.446 (0.164) | 2.024 | 0.056 |
| SDSC1 | 0.178 (0.062) | 0.14 (0.077) | 0.172 (0.065) | 1.98 | 0.06 |
| MCC | 0.472 (0.141) | 0.377 (0.216) | 0.456 (0.159) | 1.794 | 0.087 |
| VD | 1891 (2358) | 2985 (2445) | 2075 (2397) | -1.743 | 0.094 |
| MDA | 2.536 (2.139) | 4.931 (5.84) | 2.939 (3.174) | -1.717 | 0.103 |
| SDSC2 | 0.672 (0.167) | 0.553 (0.284) | 0.652 (0.195) | 1.708 | 0.103 |
| RVD | 0.085 (0.456) | 0.383 (0.902) | 0.135 (0.562) | -1.364 | 0.188 |
| Sensitivity | 0.492 (0.187) | 0.458 (0.307) | 0.487 (0.211) | 0.457 | 0.652 |
| HD95 | 14.01 (8.917) | 15.28 (15.74) | 14.23 (10.29) | -0.33 | 0.744 |
| APL1 | 1131 (395.4) | 1115 (575.6) | 1128 (427.8) | 0.114 | 0.909 |
| HDmax | 20.49 (10.37) | 20.36 (16.99) | 20.47 (11.64) | 0.032 | 0.974 |

Values are reported as mean (standard deviation). \* indicates $p < 0.05$ . Abbreviations: IoU: Intersection over Union; DSC: Dice Similarity Coefficient; SDSC1: Surface Dice Similarity Coefficient (1 voxel tolerance); MCC: Matthews Correlation Coefficient; SDSC2: Surface Dice Similarity Coefficient (2 voxel tolerance); APL1: Average Path Length (1 voxel tolerance); HDmax: Maximum Hausdorff Distance (mm); HD95: 95th Percentile Hausdorff Distance (mm); RVD: Relative Volume Difference; MDA: Mean Distance to Agreement (mm); VD: Volume Difference; AVD: Absolute Volume Difference.

The heterogeneity of geometric performance is visualized in Figure 1, where waterfall plots illustrate a tail of poor□agreement cases concentrated in the Minor Deviation/Unacceptable category. Dice, IoU, and both surface□Dice scores span from near□zero to >0.8. The 4 worst performing cases, all protocol deviations, cluster at the left of these panels. The precision and sensitivity plots exhibit similar drop-offs in these outliers, indicating possible human under□segmentation and over-segmentation respectively. In error□based metrics (MDA, VD/RVD, AVD, APL□) and distance metrics (HD□□, HD□□□), a right□hand tail is formed consisting of Minor Deviation/Unacceptable with errors several□fold higher than the median Per□Protocol case.

**Figure 1.**
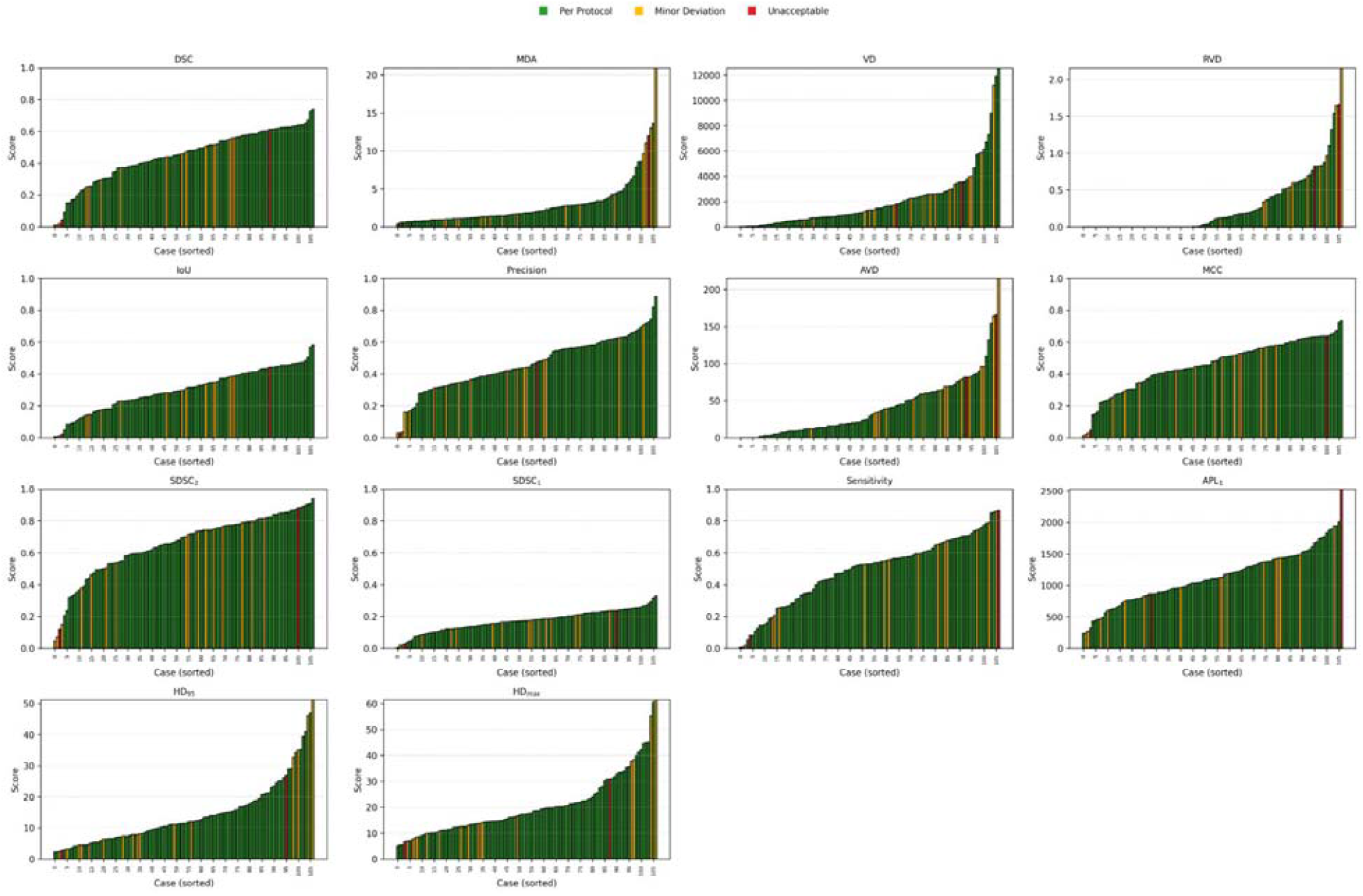
Waterfall plots of individual patient case scores for metrics describing differences between AI and human IPA segmentation

Qualitative inspection confirmed that these outliers frequently involved under- or over-segmented vascular anatomy (Figure 2). In the Per□Protocol example (Row A), the human contour coincides with the AI prediction across inferior, middle, and superior slices. In the Minor Deviation example (Row B), the human contour is under□drawn compared to the AI contour, inferiorly stopping short anteromedially and superiorly cutting off prematurely. The AI contour fills these missed segments, producing the low□precision/high□error metrics that flagged this case as not Per□Protocol.

**Figure 2.**
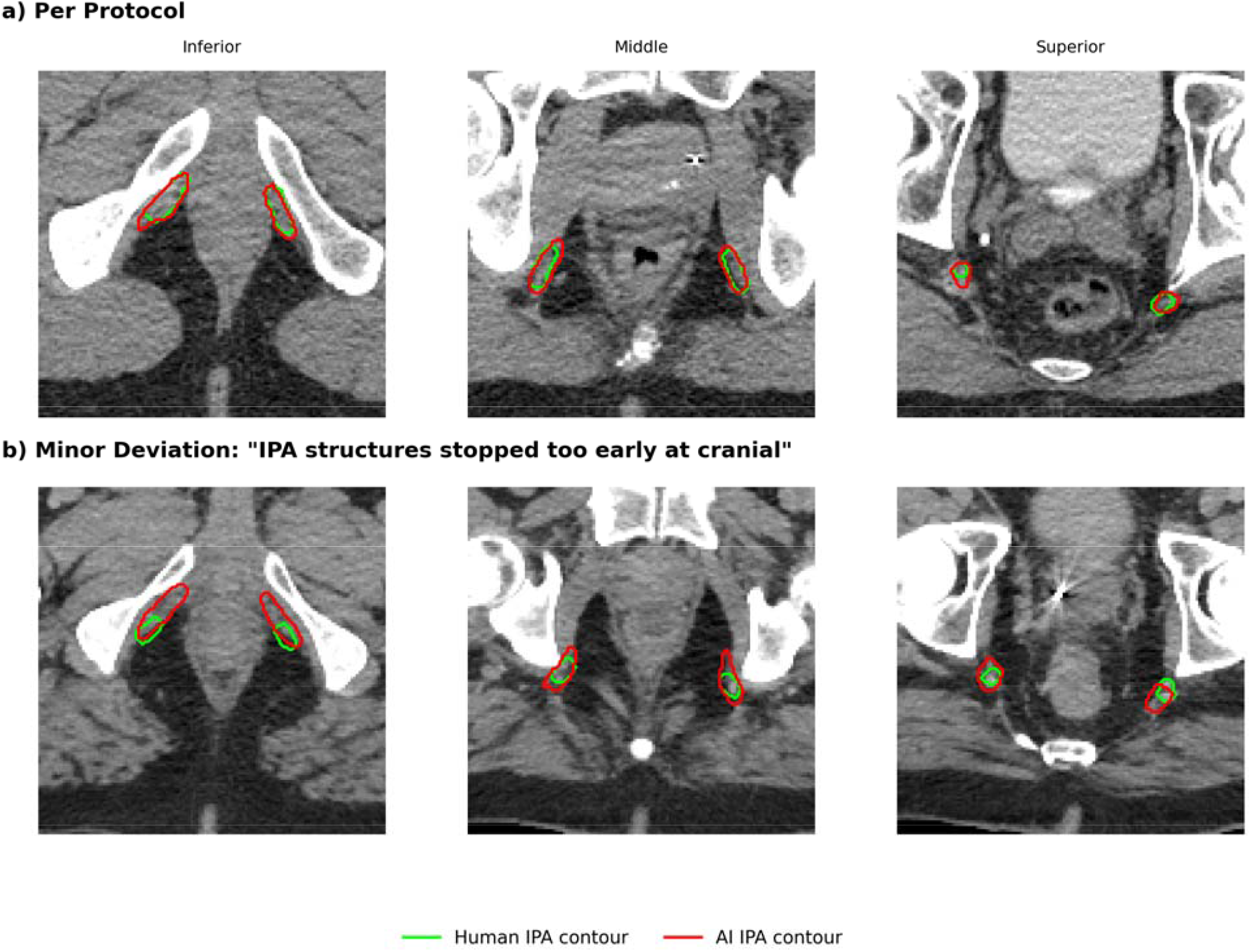
Representative comparison of Per Protocol and Minor Deviation patient cases at various anatomical positions.

### Machine□learning triage of protocol deviations

A false negative case is defined as a Minor Deviation/Unacceptable case that was not flagged by the triage classifier, while a false positive case is defined as a Per-Protocol case that was incorrectly flagged by the model. A threshold was chosen for each of the five classifiers such that all the Minor Deviation/Unacceptable cases in the training set were flagged. This strategy was used in the holdout test set with each of the five classifiers maintaining a false negative rate of 0, signifying that no deviating cases were missed across all models. The radial□basis□function SVM yielded the fewest cross-validated false positives and the highest test discrimination (AUROC = 0.836) with 6 false positive cases in the holdout set (Figure 3). Combining the cross-validation and test set cases, the SVM achieved accuracy, sensitivity, and specificity of 0.738, 1.0, and 0.685 respectively. The other classifiers underperformed the SVM overall, with logistic regression coming closest but showing a lower test AUROC of 0.818 while matching the SVM’s 6 test false positives. Performance for the remaining models was slightly weaker, with lower test AUROCs (random forest 0.718, XGBoost 0.818, feed-forward network 0.809) and more test false positives ranging from 8 to 9 (Supplemental Table 1).

**Figure 3.**
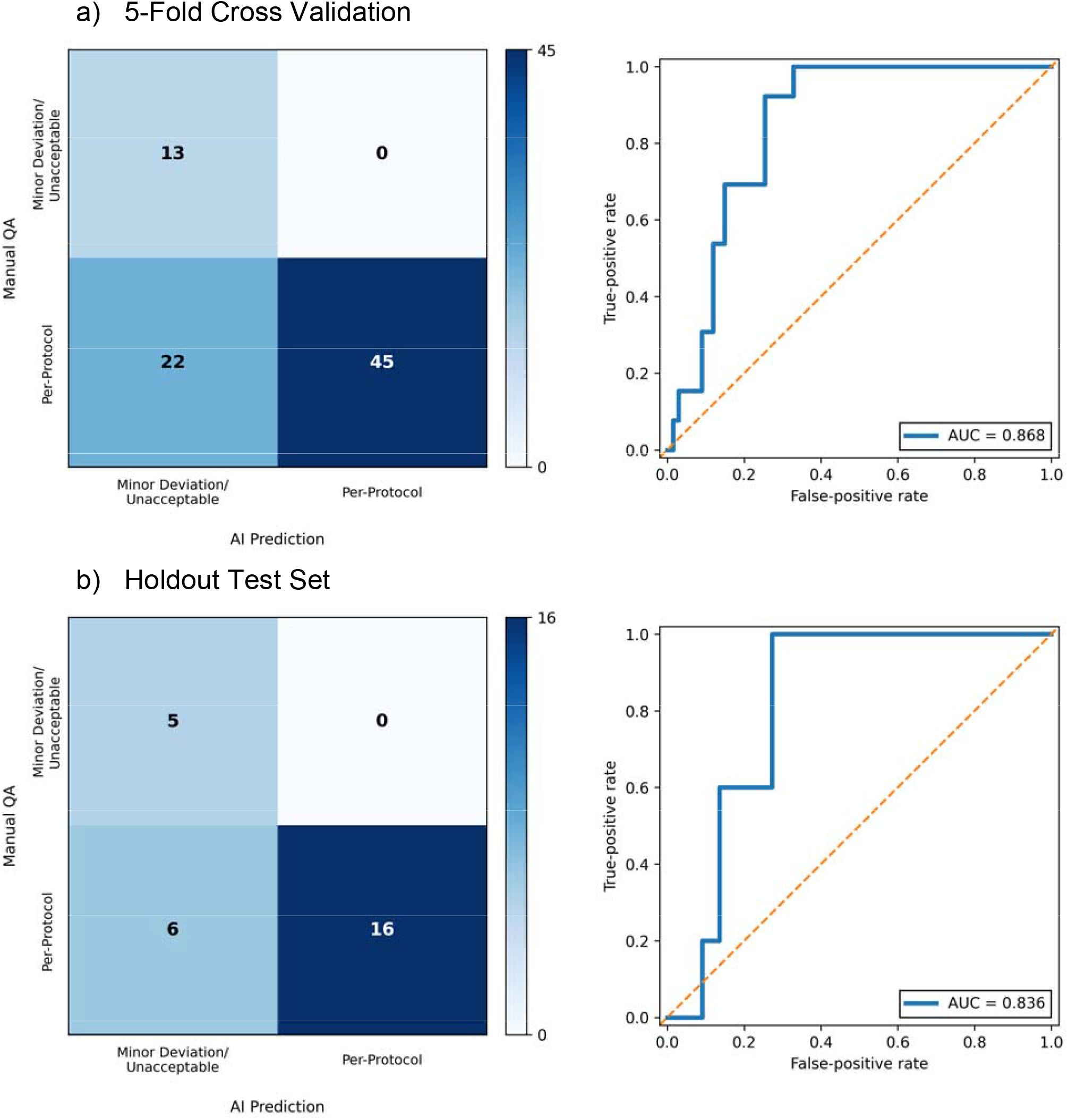
Confusion matrices and receiver-operating characteristics curves showing performance of the SVM machine learning classifier in identifying protocol deviations.

## Discussion

Our study validates a novel deep learning network for segmentation of the IPA organ-at-risk for prostate radiotherapy. Furthermore, when coupled with a SVM machine learning classifier, the AI pipeline can triage contour quality in multi-institutional clinical□trial data, identifying protocol deviations with high sensitivity and potentially augmenting expert clinician QA through decreasing the manual review burden.

The geometric performance of our AI pipeline was modest, with Dice similarity coefficient of 0.446 for the IPA, lower than typical values reported for larger pelvic organs at risk. However, this is not unexpected given the IPA’s small structure and notable inter-observer variability ^[47]^. In addition, small□volume vascular structures amplify surface discrepancies, explaining the wider Hausdorff tails we recorded. For these reasons, no single metric could be relied on to discriminate between Per-Protocol and Minor Deviation/Unacceptable cases. Nevertheless, our expanded set of 14 metrics was sufficient for using a SVM model to distinguish per protocol from deviant contours, even despite overlaps in each individual metric. Notably, relative to manual review, our study’s workflow combining AI segmentation with machine learning classification would have potentially reduced expert review workload by 57% (46 cases to review instead of 107) while still capturing all Minor Deviation/Unacceptable contours with a sensitivity of 100%. Our classifier’s AUROC of 0.836 on the independent hold-out set is in line with previous contour QA models. Duan□et□al. reported AUROCs of 0.89 for nodal lymph-node contours and 0.87 for bladder contours in a multicenter pelvic cohort, and Zhao□et□al. achieved 0.82 for lung and heart contours using a one-class SVM. These findings across pelvic and thoracic sites suggests the generalizability of contour-QA approaches to other disease sites ^[48,49]^.

Within clinical trials, radiotherapy protocol deviations have been associated with inferior patient outcomes, with contemporary and earlier meta-analyses suggesting that suboptimal radiotherapy plan quality may be associated with worse survival and disease control ^[4,50]^. In prostate cancer specifically, protocol non-adherence to contouring and planning requirements was associated with higher risk of genitourinary toxicity ^[5]^. Such deviations also threaten internal validity by introducing treatment heterogeneity that can dilute the treatment effect and make it harder to attribute observed outcomes to the intended intervention rather than radiotherapy quality ^[4,6,51]^. Multicenter trials have documented treatment planning variations and attempted use of centralized QA to reduce them ^[52]^. Because current radiotherapy QA processes are time-consuming and resource-intensive, our proposed automated IPA segmentation and SVM triage pipeline could flag potential deviations and prioritize expert review to reduce such requirements ^[53]^.

Our study has several limitations. While using multi-institutional data, this retrospective analysis used a relatively small cohort of patients with 83% Per□Protocol and 17% Minor Deviation/Unacceptable cases, and thresholds were retrospectively tuned to favorably balance false positives and false negatives which may not generalize despite nested cross□validation.

Geometry□based scores such as Dice and Hausdorff are size□dependent and may not reflect clinical outcomes, which is of particular concern for small structures like the IPA ^[25]^. Training of the IPA segmentation model occurred at a single center and required a physician□defined centroid. Differences in CT/MRI availability, scanner settings, and clinical workflow across institutions (including shifts in practice patterns over time) may introduce domain shift and erode performance, potentially contributing to lower model accuracy than was reported in the original study ^[21]^. We did not conduct expert review to assess clinical acceptability of AI-generated contours, or the degree of editing that would be required to create clinically acceptable contours, and our geometric agreement metrics alone do not establish clinical readiness of the model. We also did not evaluate QA strategies that leverage uncertainty or network□score metrics ^[54,55]^. Finally, we did not perform prospective validation or deploy the pipeline in clinical workflows, therefore any anticipated potential reduction in manual QA burden may not translate into practice without future prospective clinical evaluations.

## Conclusions

Automated segmentation of the IPA, paired with comprehensive metrics□driven machine learning triage, correctly identified all protocol deviations in a seven-center prostate SABR trial while potentially halving human expert review burden. Prospective evaluation and expansion to additional structures and clinical settings are warranted towards real□time, AI□assisted radiotherapy QA to improve both clinical trial conduct and real-world patient care delivery.

## Data Availability

The datasets analyzed in this study include de-identified clinical imaging, structure contours, and treatment-planning data generated within the POTEN-C trial and are not publicly available due to patient privacy considerations and institutional/ethical restrictions. De-identified derived data supporting the findings of this study are available from the corresponding author upon reasonable request and subject to applicable approvals and data use agreements.

## References

[1] Jomy, J. et al. Clinical impact of radiotherapy quality assurance results in contemporary cancer trials: a systematic review and meta-analysis. Radiother. Oncol. J. Eur. Soc. Ther. Radiol. Oncol. 207, 110875 (2025).

[2] Corrigan, K. L. et al. The Radiotherapy Quality Assurance Gap Among Phase III Cancer Clinical Trials. Radiother. Oncol. J. Eur. Soc. Ther. Radiol. Oncol. 166, 51–57 (2022).

[3] Smith, K. et al. Quality improvements in radiation oncology clinical trials. Front. Oncol. 13, (2023).

[4] Ohri, N. et al. Radiotherapy protocol deviations and clinical outcomes: a meta-analysis of cooperative group clinical trials. J. Natl. Cancer Inst. 105, 387–393 (2013).

[5] Beck, M. et al. Adherence to Contouring and Treatment Planning Requirements Within a Multicentric Trial: Results of the Quality Assurance of the SAKK 09/10 trial. Int. J. Radiat. Oncol. Biol. Phys. 113, 80–91 (2022).

[6] Brooks, C., Miles, E. & Hoskin, P. J. Radiotherapy trial quality assurance processes: a systematic review. Lancet Oncol. 25, e104–e113 (2024).

[7] Tree, A. C. et al. Intensity-modulated radiotherapy versus stereotactic body radiotherapy for prostate cancer (PACE-B): 2-year toxicity results from an open-label, randomised, phase 3, non-inferiority trial. Lancet Oncol. 23, 1308–1320 (2022).

[8] Tree, A. C. et al. Intensity Modulated Moderately Hypofractionationated Radiotherapy Versus Stereotactic Body Radiotherapy for Prostate Cancer (PACE-C): Early Toxicity Results From a Randomised Open-label Phase III Non-inferiority Trial. Lancet Oncol. 26, 936–947 (2025).

[9] Achard, V. et al. Urethra-Sparing Prostate Cancer Stereotactic Body Radiation Therapy: Sexual Function and Radiation Dose to the Penile Bulb, the Crura, and the Internal Pudendal Arteries From a Randomized Phase 2 Trial. Int. J. Radiat. Oncol. Biol. Phys. 119, 1137–1146 (2024).

[10] Brand, V. J. et al. Challenges and opportunities to minimize the dose in the neurovascular bundles during prostate radiotherapy. Clin. Transl. Radiat. Oncol. 53, 100959 (2025).

[11] Guevelou, J. L. et al. Sexual Structure Sparing for Prostate Cancer Radiotherapy: A Systematic Review. Eur. Urol. Oncol. 7, 332–343 (2024).

[12] Kwakernaak, R. C. et al. Neurovascular bundle sparing in hypofractionated radiotherapy maintained with realistic treatment uncertainties. Phys. Imaging Radiat. Oncol. 33, (2025).

[13] Desai, N. B. et al. Prostate oncologic therapy while ensuring neurovascular conservation (POTEN-C): A phase II randomized controlled trial of stereotactic ablative body radiotherapy (SAbR) with or without neurovascular sparing for erectile function preservation in localized prostate cancer. J. Clin. Oncol. 38, TPS381–TPS381 (2020).

[14] POTEN-C Trial. https://www.utsouthwestern.edu/departments/radiation-oncology/research/clinical/poten-c/index.html.

[15] Pang, E. P. P. et al. Multicentre evaluation of deep learning CT autosegmentation of the head and neck region for radiotherapy. NPJ Digit. Med. 8, 312 (2025).

[16] Kehayias, C. E. et al. Prospective deployment of an automated implementation solution for artificial intelligence translation to clinical radiation oncology. Front. Oncol. 13, 1305511 (2024).

[17] Matoska, T., Patel, M., Liu, H. & Beriwal, S. Review of Deep Learning Based Autosegmentation for Clinical Target Volume: Current Status and Future Directions. Adv. Radiat. Oncol. 9, (2024).

[18] Kibudde, S. et al. Impact of Artificial Intelligence-Based Autosegmentation of Organs at Risk in Low- and Middle-Income Countries. Adv. Radiat. Oncol. 9, (2024).

[19] Rong, Y. et al. NRG Oncology Assessment of Artificial Intelligence Deep Learning-Based Auto-segmentation for Radiation Therapy: Current Developments, Clinical Considerations, and Future Directions. Int. J. Radiat. Oncol. Biol. Phys. 119, 261–280 (2024).

[20] Moran, K., Poole, C. & Barrett, S. Evaluating deep learning auto-contouring for lung radiation therapy: A review of accuracy, variability, efficiency and dose, in target volumes and organs at risk. Phys. Imaging Radiat. Oncol. 33, 100736 (2025).

[21] Balagopal, A. et al. Deep learning based automatic segmentation of the Internal Pudendal Artery in definitive radiotherapy treatment planning of localized prostate cancer. Phys. Imaging Radiat. Oncol. 30, 100577 (2024).

[22] Poel, R. et al. A comprehensive multifaceted technical evaluation framework for implementation of auto-segmentation models in radiotherapy. Commun. Med. 5, 319 (2025).

[23] Desai, N. A Phase II Randomized Controlled Trial of Stereotactic Ablative Body Radiotherapy (SAbR) With or Without Neurovascular Sparing for Erectile Function Preservation in Localized Prostate Cancer. https://clinicaltrials.gov/study/NCT03525262 (2025).

[24] Vaassen, F. et al. Evaluation of measures for assessing time-saving of automatic organ-at-risk segmentation in radiotherapy. Phys. Imaging Radiat. Oncol. 13, 1–6 (2019).

[25] Sherer, M. V. et al. Metrics to evaluate the performance of auto-segmentation for radiation treatment planning: A critical review. Radiother. Oncol. J. Eur. Soc. Ther. Radiol. Oncol. 160, 185–191 (2021).

[26] Harrison, K. et al. Machine Learning for Auto-Segmentation in Radiotherapy Planning. Clin. Oncol. 34, 74–88 (2022).

[27] Mackay, K., Bernstein, D., Glocker, B., Kamnitsas, K. & Taylor, A. A Review of the Metrics Used to Assess Auto-Contouring Systems in Radiotherapy. Clin. Oncol. 35, 354–369 (2023).

[28] Sanders, J. W. et al. Prospective Evaluation of Prostate and Organs at Risk Segmentation Software for MRI-based Prostate Radiation Therapy. Radiol. Artif. Intell. 4, e210151 (2022).

[29] Xue, X. et al. Deep learning-based segmentation for high-dose-rate brachytherapy in cervical cancer using 3D Prompt-ResUNet. Phys. Med. Biol. 69, (2024).

[30] Nai, Y.-H. et al. Comparison of metrics for the evaluation of medical segmentations using prostate MRI dataset. Comput. Biol. Med. 134, 104497 (2021).

[31] Urago, Y. et al. Evaluation of auto-segmentation accuracy of cloud-based artificial intelligence and atlas-based models. Radiat. Oncol. 16, 175 (2021).

[32] Zhu, J. et al. Evaluation of Automatic Segmentation Model With Dosimetric Metrics for Radiotherapy of Esophageal Cancer. Front. Oncol. 10, (2020).

[33] Cha, E. et al. Clinical implementation of deep learning contour autosegmentation for prostate radiotherapy. Radiother. Oncol. J. Eur. Soc. Ther. Radiol. Oncol. 159, 1–7 (2021).

[34] Cortes, C. & Vapnik, V. Support-vector networks. Mach. Learn. 20, 273–297 (1995).

[35] Chen, J., Phan, T. G. & Reutens, D. C. Ridge penalized logistical and ordinal partial least squares regression for predicting stroke deficit from infarct topography. J. Biomed. Sci. Eng. 3, 568–575 (2010).

[36] Breiman, L. Random Forests. Mach. Learn. 45, 5–32 (2001).

[37] Ishwaran, H. & O’Brien, R. Commentary: The Problem of Class Imbalance in Biomedical Data. J. Thorac. Cardiovasc. Surg. 161, 1940–1941 (2021).

[38] Leevy, J. L., Khoshgoftaar, T. M., Bauder, R. A. & Seliya, N. A survey on addressing high-class imbalance in big data. J. Big Data 5, 42 (2018).

[39] Chen, T. & Guestrin, C. XGBoost: A Scalable Tree Boosting System. in Proceedings of the 22nd ACM SIGKDD International Conference on Knowledge Discovery and Data Mining 785–794 (Association for Computing Machinery, New York, NY, USA, 2016). doi:10.1145/2939672.2939785.

[40] Lee, H., Kim, Y., Yang, S. Y. & Choi, H. Improved weight initialization for deep and narrow feedforward neural network. Neural Netw. 176, 106362 (2024).

[41] Rezvani, S., Pourpanah, F., Lim, C. P. & Wu, Q. M. J. Methods for class-imbalanced learning with support vector machines: a review and an empirical evaluation. Soft Comput. 28, 11873–11894 (2024).

[42] Dorn, M. et al. Comparison of machine learning techniques to handle imbalanced COVID-19 CBC datasets. PeerJ Comput. Sci. 7, e670 (2021).

[43] Gao, L., Liu, Z.-X. & Wang, J.-N. Predictive model and risk analysis for outcomes in diabetic foot ulcer using eXtreme Gradient Boosting algorithm and SHapley Additive exPlanation. World J. Diabetes 16, (2025).

[44] Varma, S. & Simon, R. Bias in error estimation when using cross-validation for model selection. BMC Bioinformatics 7, 91 (2006).

[45] West, R. M. Best practice in statistics: Use the Welch t-test when testing the difference between two groups. Ann. Clin. Biochem. 58, 267–269 (2021).

[46] Delacre, M., Lakens, D. & Leys, C. Why Psychologists Should by Default Use Welch’s t-test Instead of Student’s t-test. Int. Rev. Soc. Psychol. 30, (2017).

[47] Teunissen, F. R. et al. Interrater agreement of contouring of the neurovascular bundles and internal pudendal arteries in neurovascular-sparing magnetic resonance-guided radiotherapy for localized prostate cancer. Clin. Transl. Radiat. Oncol. 32, 29–34 (2021).

[48] Zhao, Y. et al. Streamlining Thoracic Radiotherapy Quality assurance: One-Class Classification for Automated OAR Contour Assessment. Technol. Cancer Res. Treat. 24, 15330338251345895 (2025).

[49] Duan, J. et al. Contouring quality assurance methodology based on multiple geometric features against deep learning auto-segmentation. Med. Phys. 50, 2715–2732 (2023).

[50] Jomy, J. et al. Clinical impact of radiotherapy quality assurance results in contemporary cancer trials: a systematic review and meta-analysis. Radiother. Oncol. 207, (2025).

[51] Corrigan, K. L. et al. The Radiotherapy Quality Assurance Gap Among Phase III Cancer Clinical Trials. Radiother. Oncol. J. Eur. Soc. Ther. Radiol. Oncol. 166, 51–57 (2022).

[52] Kearvell, R. et al. Quality improvements in prostate radiotherapy: outcomes and impact of comprehensive quality assurance during the TROG 03.04 ‘RADAR’ trial. J. Med. Imaging Radiat. Oncol. 57, 247–257 (2013).

[53] Nielsen, C. P. et al. Recommendations for radiotherapy quality assurance in clinical trials. Radiother. Oncol. 209, (2025).

[54] Wahid, K. A. et al. Harnessing uncertainty in radiotherapy auto-segmentation quality assurance. Phys. Imaging Radiat. Oncol. 29, 100526 (2024).

[55] Rodríguez Outeiral, R. et al. A network score-based metric to optimize the quality assurance of automatic radiotherapy target segmentations. Phys. Imaging Radiat. Oncol. 28, 100500 (2023).

